# A Mendelian randomization study of telomere length and blood-cell traits

**DOI:** 10.1101/2020.05.30.20117978

**Authors:** Charleen D. Adams, Brian B. Boutwell

## Abstract

Whether telomere attrition reducing proliferative reserve in blood-cell progenitors is causal has important public-health implications. Mendelian randomization (MR) is an analytic technique using germline genetic variants as instrumental variables. If certain assumptions are met, estimates from MR should be free from most environmental sources of confounding and reverse causation. Here, two-sample MR is performed to test whether longer telomeres cause changes to hematological traits. Summary statistics for genetic variants strongly associated with telomere length were extracted from a genome-wide association (GWA) study for telomere length in individuals of European ancestry (n=9190) and from GWA studies of blood-cell traits, also in those of European ancestry (n∼173,000 participants). A standard deviation increase in genetically influenced telomere length increased red blood cell (RBC) and white blood cell (WBC) counts, decreased mean corpuscular hemoglobin (MCH) and mean cell volume (MCV), and had no observable impact on mean corpuscular hemoglobin concentration (MCHC), red cell distribution width (RDW), hematocrit, or hemoglobin. Sensitivity tests for pleiotropic distortion were mostly inconsistent with glaring violations to the MR assumptions. Similar to how germline mutations in *TERT* can lead to bone-marrow failure, these data provide evidence that genetically influenced common variation in telomere length impacts hematologic traits in the population.

---

In humans, telomeres, DNA repeats and protein structures that “cap” and stabilize the ends of chromosomes, progressively shorten across the lifespan^1^. As a result, telomere length has generally been viewed as a biomarker of aging^2^. Natural variation for telomere length exists, moreover, with a portion of the variance resulting from genetic variation^3^ (e.g., heritability estimates ranging from 36 – 82%^4^). While heritable influences likely capture a variety of contributing mechanisms, there is reason to suspect that they heavily reflect both the activity of the ribonucleoprotein enzyme telomerase, which acts to maintain telomere length, as well as the effects of environmental (e.g. exogenous oxidative or inflammatory) stressors that can accelerate telomere shortening^5,6^.

Blood, like telomere length, affects and is affected by its environment: what happens in and to blood impacts physiological processes throughout the body, and blood cells also carry the signatures of environmental stressors (e.g., smoking- and alcohol-related DNA methylation changes in leukocytes^7,8^). Telomere length can be thought of as an environmentally pliant and endogenous exposure for cells. A DNA damage response may be elicited and cellular senescence ensue if telomeres become critically short^9^. Telomere length in white blood cells (WBCs) may approximately reflect telomerase activity in hematopoietic stem cells (HSCs)^10^. This becomes broadly relevant, to be sure, as telomerase activity appears to be inadequate at preventing telomere erosion^11^. As hypothesized by Mazidi, Penson, and Banach (2017), telomere attrition may be a marker for reduced proliferative reserve in hematopoietic progenitor cells^12^.

An established relationship exists between rare telomerase mutations and bone marrow failure syndromes^13^. As Savage and Bertuch (2010) discuss in their review, telomere biology disorders are, indeed, defined by very short telomeres. Affected persons are highly susceptible to cancer, pulmonary fibrosis, and bone marrow failure (BMF)^4^. BMF can be the first sign of a telomere biology disorder, in fact. Aplastic anemia, a BMF condition which can be acquired or inherited and which results from damage to HSCs^14^, leads to hallmark cytopenia: low circulating RBCs, WBCs, and platelets. Fascinatingly, telomere length can be shorter than normal in those with acquired aplastic anemia, but not as short as for those with canonical telomere biology disorders^4^. Likewise, lower red blood cell (RBC) counts, larger RBC size, and lower platelet counts have been observed in subjects with germline telomerase reverse transcriptase (*TERT*) mutations compared with family controls^15^.

At least 70% of patients with dyskeratosis congenita, which has the most severe phenotype of the telomere biology disorders^4^, carry germline mutations in telomere maintenance genes^13^. Moreover, similar to telomere length in those with acquired anemia sometimes being shorter than in non-affected individuals—but not as short as in those with established telomere biology disorders^4^—telomere length in those with inherited bone marrow failure syndromes (IBMFSs) other than dyskeratosis congenita have been documented to be shorter than in unaffected individuals. But they are not as short as in those dyskeratosis congenita^13^. This implies a dose-like relationship between the heritable component of telomere length and disease severity, which is especially notable with BMF in dyskeratosis congenita, associated with accelerated telomere shortening^16^.

The heritable component to telomere length may not necessarily be limited to mutations directly in telomere biology, however. For instance, there is a lack of evidence for mutations in telomere biology genes in patients with Fanconi anemia (an IBMFS), for which BMF is the most likely adverse outcome during childhood (relative to solid tumors being more likely in adulthood), and in which shorter telomeres have been documented^17^. With Fanconi anemia, Pang and Andreassen (2009) and Sarkar and Liu (2016) postulate that telomere defects occur secondary to endogenous oxidative damage^18,19^.

By extension, these hints of dose-like relationships raise the possibility that common variation (like a small dose in susceptibility from a single nucleotide polymorphism [SNP] rather than a large dose from a rare mutation) in telomere maintenance genes might confer an effect on blood-cell traits, though in a blunted, ostensibly subclinical, manner in comparison to the mutations causing IBMFS. And, while subclinical, these changes could still be relevant to the pathophysiology of common diseases of aging in the population.

Telomere length is, indeed, hypothesized to be associated with hematologic traits in the general population. For instance, in an observational study of 3156 subjects, Kozlitina *et al*. (2012) found that shorter telomere lengths were associated with lower RBC counts, larger mean RBC size, increased red blood cell distribution width (RDW), higher hemoglobin levels, and lower platelet counts^6^.

The Kozlitina study was small, however, and observational studies are prone to potential confounding and reverse causation. Indeed, a few other small studies have examined telomere length and blood-cell traits with results that are discrepant between the studies^12^. To wit, the results for the study by Mazidi, Penson, and Banach (2017) partly conflict with those by Kozlitina *et al*. (2012). They observed a negative relationship between telomere length and monocyte count, for instance^12^. Due to this, it is unclear whether a causal relationship exists between telomere length and blood-cell traits in the general population.

Using Mendelian randomization (MR), Haycock *et al*. (2017) observed that longer telomeres (in leukocytes) increased the risk for various cancer outcomes, but decreased the risk for coronary heart disease^20^. Since variation in blood-cell subtype is associated with a wide variety of systemic diseases^21^ and since blood cells play an essential role in key physiological processes (e.g., oxygen transport, hemostasis, and innate and acquired immune responses)^22–24^, the activity of blood-cell traits may mediate or exacerbate some of the effects of telomere length on various disease outcomes and/or reflect changes resulting from various diseases. Teasing out the underlying causal relationships has important clinical and public-health implications.

To that aim, MR is an analytic, quasi-experimental technique that uses germline genetic variants as instrumental variables. If certain assumptions are met, estimates from MR should be free from most social and environmental sources of confounding and reverse causation (see Methods)^25^, and can, therefore, help sort out the nature of complexly woven traits. Here, MR is used to examine the effect of genetically influenced longer telomeres on nine blood-cell traits.

## Results

### Red blood cell (RBC) and white blood cell (WBC) counts

Genetically increased telomere length was associated with higher inverse-variance weighted (IVW) estimates (95% CIs) for 2 of 9 blood-cell traits (*P* < 0.006): RBC count 0.09 (0.04, 0.14) and WBC count 0.06 (0.02, 0.11). The sensitivity estimators aligned in the direction and magnitudes of their effects, and the MREgger intercept tests (Supplementary Tables 10–11) indicated no evidence for directional pleiotropy. The *I*^2^ statistic for the MR-Egger test pointed to some potential regression dilution, though the values were close to 90%, indicating that the MR-Egger tests suffered at most from up to 14% potential dilution from this bias. The Simulation Extraction (SIMEX) correction of the MR-Egger estimates revealed that the corrected MR-Egger intercepts were different than zero (an MR-Egger intercept test consistent with zero indicates a lack of evidence for pleiotropy in the IVW estimate). While this could imply distortion from pleiotropy in the IVW estimate (Supplementary Tables 10–11), the SIMEX correction may be overly conservative, since the potential dilution was small.

### Reticulocyte count

There was intermediate evidence that genetically increased telomere length was associated with higher reticulocyte count (*P* < 0.05): IVW estimate 0.04 (95% CI 0.00, 0.08). The sensitivity estimators aligned in the direction and magnitudes of their effects, except, crucially, for the MR-Egger estimate, for which the direction of effect was reversed. The MREgger intercept test (Supplementary Table 12) indicated no evidence for directional pleiotropy. The *I*^2^ statistic for the MR-Egger test pointed to some potential regression dilution, indicating that the MR-Egger test suffered from up to 13% potential dilution from this bias (potential bias of <10% is typically considered acceptable). The SIMEX correction of the MR-Egger estimate made no difference in the interpretation of the findings (the MR-Egger intercept remained null). Overall, however, due to the discordance between the IVW and the MR-Egger estimates, the finding could be influenced by violations to the MR assumption against horizontal pleiotropy.

### Mean corpuscular hemoglobin (MCH) and mean corpuscular (cell) volume (MCV)

Genetically increased telomere length was associated at the Bonferroni threshold with lower IVW estimates [95% CIs] for 2 of the 9 blood-cell traits (*P* < 0.006): MCH (−0.12 [−0.18, –0.07]) and MCV (−0.13 [−0.18, –0.08]). The sensitivity estimators aligned in their direction of effects and mostly in their magnitudes. The MR-Egger intercept tests (Supplementary Tables 13–14) indicated no evidence for directional pleiotropy. The *I*^2^ statistic for the MR-Egger test to minimal (up to 11%) potential dilution. SIMEX correction of the MR-Egger estimates indicated that the corrected MR-Egger intercepts remained consistent with zero after correction (Supplementary Tables 13–14).

### Mean corpuscular hemoglobin concentration (MCHC), red cell distribution width (RDW), hematocrit (Hct), and hemoglobin (Hgb)

Genetically increased telomere length was not associated with 4 of the 9 blood-cell traits: MCHC (−0.02 [−0.06, 0.02]), RDW (0.02 [−0.02, 0.06]), Hct (0.03 [−0.02, 0.08]), RDW (0.04 [0.00, 0.08]). The sensitivity estimators mostly aligned in their magnitudes of effects, but for the MCHC estimate, several of the estimators were reversed in direction from that of the IVW. The MR-Egger intercept tests (Supplementary Tables 15–18) indicated no evidence for directional pleiotropy. The *I*^2^ statistic for the MR-Egger test indicated up to ∼15% potential dilution from this bias. The SIMEX correction of the MR-Egger estimates indicated that the corrected MR-Egger intercepts remained consistent with zero after correction (Supplementary Tables 15–18): no change in interpretation.

## Discussion

### Summary

The MR analysis of telomere length on blood-cell traits support the notion that telomere length influences some of the traits in a causal manner. Longer telomeres increased RBC and WBC counts and decreased the measures of MCH and MCV, but had no observable impact on MCHC, RDW, hematocrit, or hemoglobin. The sensitivity tests for potential pleiotropic distortion were mostly inconsistent with glaring violations to the MR assumptions. Supposing the MR assumptions have not been violated, which can never fully be tested (a limitation of this study and of all MR studies generally), these data support the growing evidence that longer telomeres impact blood cells.

### Relevance for disease and aging

As for what this means for the clinic and for the population at large, De Meyer *et al*. (2008) suggested that there may be an age-dependent effect of telomere length on blood-cell traits. In their population-based study, they observed a positive correlation between telomere length and RBC count that was, importantly, stronger for those older than 45 years of age^26^. This suggests that the impact of telomere length on RBCs may be negligible in middle age but have important clinical consequences in the elderly, related to the anemia of chronic disease^26^. Thus, longer telomeres increasing RBC count may be beneficial and protective, especially during the later years of life.

The protective effect may be dampened some, however, by the fact that no relation was observed for longer telomere length and hemoglobin concentration in the present study. Nonetheless, a pattern of longer telomeres increasing RBCs and WBCs but not hemoglobin concentration comports with those of an observational study among Polish individuals over 65^10^. For men specifically, Gutmajster *et al*. (2013) detected weak positive correlations between telomere length and RBCs and WBCs and no signal for hemoglobin. The present data dovetails with theirs, supporting the hypothesis that telomere shortening hinders hematopoiesis capacity.

Moreover, in the Kozlitina *et al*. (2012) study, an inverse association was observed between telomere length and MCV^6^. Our findings for MCV comport with Kozlitina *et al*.’s and support the idea that telomere shortening is a mechanism for the macrocytosis of aging.

Haycock *et al*. (2017) observed that increased telomere length increased the odds for lung adenocarcinoma (odds ratio and 95% CI: 3.19 [2.40–4.22]) and other cancers^20^, and Sprague *et al*. (2009) documented that higher WBC count increased the hazard estimates for lung cancer^27^. Thus, it is conceivable that there is a pathway from longer telomeres to lung cancer through an impact on increasing WBC count, a marker of inflammation^28^. A body of evidence suggests that chronic low-grade inflammation is involved in the pathogenesis of various cancers, where cancer causes inflammatory, microenvironmental, and immune changes to the host^29–33^. Therefore, since longer telomeres appear to contribute to the initiation of some cancers and these early carcinogenic changes potentially promote increased WBC count, increased WBC count might reflect undiagnosed cancer. If longer telomeres increase WBC counts and contribute to cancer initiation, and, in turn, the subsequent (possibly subclinical) tumor increases WBC, this would be an example of bidirectional causality.

### Bidirectional causality versus reversion causation

Bidirectional causality is different conceptually from reverse causation, and because it could be relevant to our findings, it is useful to draw out the distinctions. With a true bidirectional relationship, causation occurs in both directions, as the name suggests. For example, there is a bidirectionally causal relationship between fluid intelligence and years of schooling. Higher intelligence causes people to stay in school longer, and staying in school longer feeds back on and increases intelligence^34^. This bidirectionality is different than what is typically meant by “reverse causation”. In non-experimental studies, where temporality cannot be confidently sussed out, the direction of the correlation is challenging, or impossible, to infer. When the actual underlying causal direction is opposite from what investigator’s hypothesize, this is an example of reverse causation. MR studies are specifically designed to subvert reverse causation. This is so, owing to the fact that genotype assignment at conception temporarily precedes other physiological parameters of interest. This robustness to reverse causality is one of the overarching strengths of MR and why MR is an essential tool for population-based medical research. Relative to observational designs, the risk for reverse causation is greatly reduced.

However, it is not impossible, as VanderWeele *et al*. (2014) point out. Scenarios can arise in which an exposure and outcome of interest are each partitioned into several time points. For instance, imagine an outcome at its “time 1” that affects an exposure at the exposure’s “time 2”. This can approximate reverse causation in MR since the genetic variant proxying for the exposure at “time 1” is now not independent of the final outcome at “time 2”, conditional on both exposure time points and potential confounders^35^. Fortunately, this should be the exception rather than the rule. While possible that one exists, absent knowledge of an empirical, a prior reason to think that telomeres impact blood-traits at multiple time points in such a way that blood-cell traits influence telomeres at “time 2”, a more parsimonious explanation is that the relationships between telomeres and blood cells are bidirectional, if indeed blood-cell traits impact telomeres. Blood-cell traits might very well do so, given that blood-cell traits are involved in oxidative and inflammatory processes.

Whether blood-cell traits cause changes in telomere length is an important avenue for future MR research. In order to examine the influence of blood-cell traits on telomere length with two-sample MR (the design used here), the full summary statistics for a *large* telomere-length GWA studies would be needed. While the findings deemed “statistically significant” for the telomere-length GWA study used here to instrument telomere length are public, the full summary statistics, including the non-top findings, are not available. For now, this precludes MR investigations of bidirectional relationships between longer telomeres and blood-cell traits using the Mangino *et al*. GWA data as the outcome data source. Additionally, and more importantly, a telomere-length GWA study that is substantially larger than Mangino *et al’s*. (2012), which included ∼9200 participants, would be needed to best capitalize on the power of large samples to detect effects. Another GWA study of telomere length does exist: Codd *et al*. (2013) contains ∼38,000 individuals^36^. This is the more-apt GWA dataset to use when treating telomere length as the outcome, (or even better, an even larger one). At present the full summary statistics are not publicly available. To further emphasize this issue, the Astle *et al*. (2016) GWA studies for blood-cell traits we used here were performed on ∼173,000 individuals. This makes the Astle *et al*. (2016) GWA studies strong resources for studying blood-cell traits as outcome variables in two-sample MR.

Returning briefly to the example of the bidirectional relationship between intelligence and education years, a bidirectional MR analysis of these traits was able to determine the prevailing direction of effect. Specifically, and intriguingly, while intelligence influences how long someone stays in school, the magnitude of the impact of staying in school on intelligence is much larger^34^. This has important implications for interventional policies. Similarly, determining the prevailing direction of effect, if a bidirectional relationship between telomere length and blood-cell traits exists, has the potential to inform and refine strategies for disease prevention.

The issue of bidirectionality gets thicker when turning attention to coronary heart disease (CHD). A potentially puzzling relationship exists between longer telomeres, more WBCs, and CHD. Haycock *et al’s*. (2017) MR study and also evidence from clinical studies suggest a protective effect of longer telomeres against CHD^20,37^. But a vast body of clinical and epidemiologic literature suggests that higher WBC counts contribute to CHD’s pathogenesis. This makes it challenging to explain the impact of telomeres increasing WBCs, as they relate to CHD. The proposed mechanisms for the deleterious effects of WBCs on CHD, as entrenched and entangled as they are with likely bidirectional causality, are hard to ignore. For instance, previously suggested “mechanistic” explanations include the fact that higher WBC counts are a biomarker of atherosclerotic stress; a secondary signal from inflammation due to tobacco smoking; a contributor to microvascular injury; and a reflection of complex and systemic inflammatory responses, involving cytokines, cell-adhesion molecules, *T*-lymphocytes, and C-reactive protein^38^, etc. But if the observational evidence is causal, then this suggests that longer telomeres and more WBCs have, perhaps, independent and opposing direct effects on CHD, even if longer telomeres increase WBC count, as suggested by the data here. Whether there are independent, direct effects of more WBCs and longer telomeres on CHD is a question that could be investigated with multivariable MR – once full summary statistics are available for telomere length (again from a large GWA study).

Lastly, chronic inflammation, independent of shorter telomere length, can lead to premature senescence. This may enhance the effects of shorter telomeres, which also cause senescence when short enough^37,39^. Moreover, since a) longer telomeres in WBCs likely reflect longer telomeres in hematopoietic stem cells (HSCs) and b) epithelial progenitor cells (EPCs) originate from HSCs and are involved in the repair mechanisms for vascular atherosclerosis^37^, shortened telomeres in WBCs may reflect compromised capacity to repair injured vasculature. Therefore, longer telomeres may be protective against CHD by not interfering with the generation of EPCs and their involvement in vasculature repair. To the extent that more WBCs reflect EPC health and integrity, more WBCs may be a protective marker. To the extent that more WBCs reflect increased inflammation, more WBCs appear to contribute to CHD.

In conclusion, similar to how germline mutations in telomere biology genes can lead to BMF, the present findings from two-sample MR provide evidence that genetically influenced common variation in telomere length impacts hematologic traits in the population. Future MR studies should explore whether blood-cell traits also impact telomere length, as similar to endogenous oxidative damage in Fanconi anemia possibly influencing telomere length, oxidative damage related to immune cell responses might influence telomeres in the population.

## Methods

### Conceptual approach

Non-experimental studies are prone to confounding and reverse causation. MR gets around these issues, using an instrumental-variables framework, by exploiting the random assortment of alleles, genotype assignment at conception, and pleiotropy (genes influencing more than one trait)^25,40,41^. Using genetic variants instrumentally avoids most environmental sources of confounding, and genotype assignment at conception avoids most sources of reverse causation, since genotypes temporarily precede the observational variables of interest.

Two-sample MR is a version of the procedure that uses summary statistics from two genome-wide association (GWA) studies^42–47^. Typically, with two-sample MR, the IVW method is the standard approach (Fig. 1 contains an example).

**Fig. 1.**
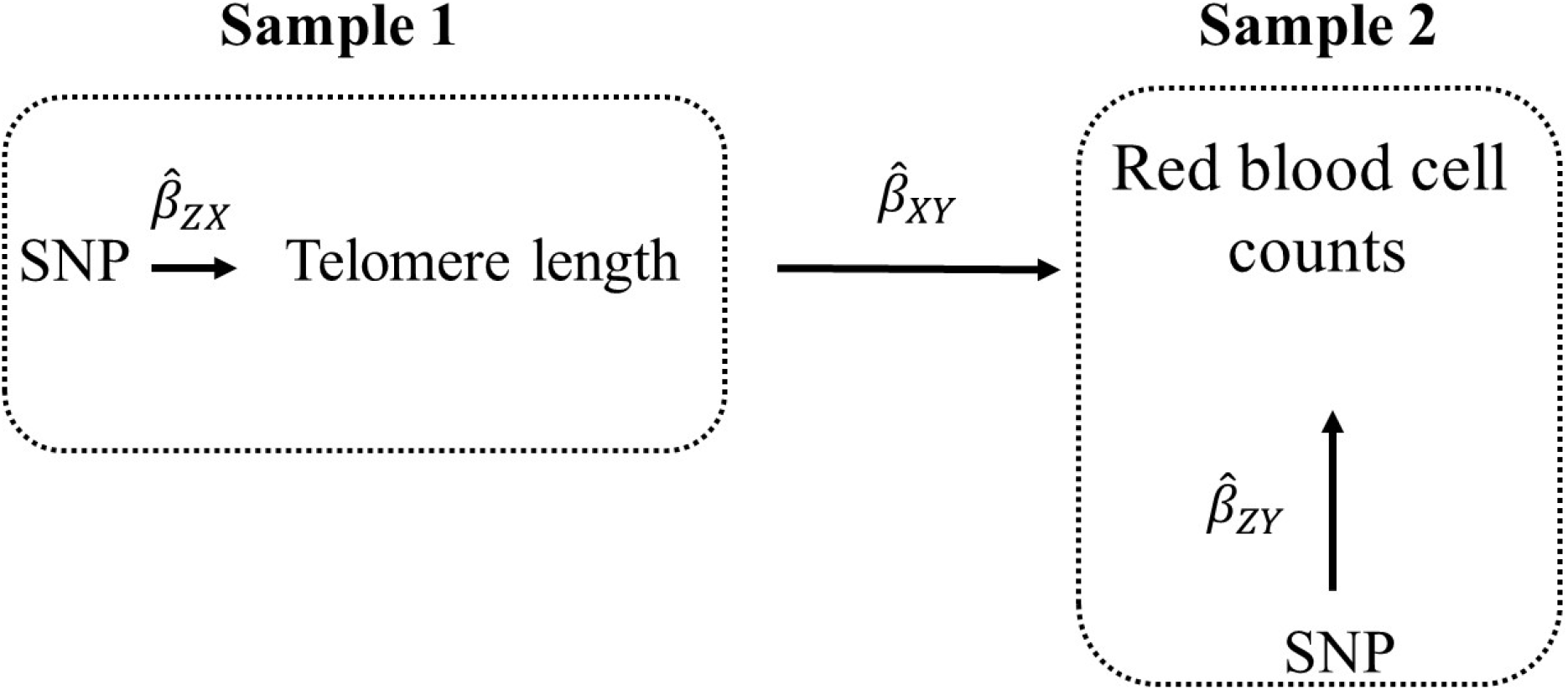
Illustration of two-sample MR using the test of the causal effect of telomere length on red blood cell (RBC) counts as an example. Estimates of the SNP-telomere length associations (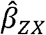) are calculated in sample 1 (from a previous GWA study of telomere length). The association between these same SNPs and RBC counts are then estimated in sample 2 (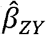) (from a GWA study of RBC counts). The estimates from the instrumental SNPs are combined into Wald ratios (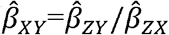). The 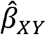 ratio estimates are meta-analyzed using the inverse-variance weighted (IVW) analysis (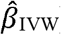) method, which produces an overall causal estimate of telomere length on RBC counts. Sensitivity estimators are used to judge whether the IVW causal estimate is plausible (more below).

### Mendelian randomization assumptions

MR relies on the validity of three assumptions^48^. In the context of the present analysis, these assumptions are as follows: (i) the SNPs acting as the instrumental variables for telomere length are strongly associated with telomere length; (ii) the telomere-length-associated SNPs are independent of confounders of telomere length and the outcomes of interest; and (iii) the telomere length SNPs are associated with the outcomes of interest only through telomere length (no horizontal pleiotropy; the SNPs are not associated with the outcomes independent of telomere length^44,48^).

### Instrument construction

For the telomere length instruments (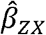 in Fig. 1), SNPs associated at genome-wide significance (*P* < 5 × 10^−8^) with a standard deviation (SD) in telomere length, whose summary statistics (effect estimates and standard errors) were concatenated and reported by Haycock *et al*. (2017)^20^ were selected. The original telomere-length GWA study was a meta-analysis performed across six cohorts on 9190 individuals (men and women aged 18 to 95) of European ancestry^3^. (Detailed descriptions of the six cohorts are available elsewhere^49–54^, but for ease of reference, Mangino *et al*. (2012) included the following six cohorts in their meta-analysis of telomere length: the Framingham Heart Study, Family Heart Study, Cardiovascular Health Study, Bogalusa Heart Study, HyperGEN, and TwinsUK). Mangino *et al*. (2012) adjusted for age, age^2^ sex, and smoking history and checked for non-European ancestry with principal components analysis. From the meta-analysis, sixteen SNPs were available for this MR analysis (Supplementary Table 19). Those that were independent (not in linkage disequilibrium, LD, with an r^2^ < 0.001, at a clumping distance of 10,000 kilobases with reference to the 1000 Genomes Project http://www.internationalgenome.org/ were kept and those that did not fit this criteria were dropped. The corresponding effect estimates and standard errors for the retained SNPs were then obtained from the blood-trait GWA studies (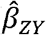 in Fig. 1) The blood-trait GWA studies were performed by Astle *et al*. (2016) on a population of ⁁173,000 individuals of European ancestry, largely from the UK (a meta-analysis of the UK Biobank and Interval studies)21. They adjusted for principal components and study center and excluded those with blood cancers or major blood disorders.

When a SNP was not available in the blood trait GWA studies, a “proxy” SNP in LD with the SNP at r^2^≥.0.80 (assessed using 1000 Genomes Project) was chosen. If the “proxy” SNP was not available, the SNP was removed from the analysis. SNP-exposure and SNP-outcome associations were harmonized with the “harmonization_data” function within the MR-Base “TwoSampleMR” package within R^42,55^. Harmonized SNP-exposure and SNP-outcome associations were combined with the IVW method (Fig. 1).

For all tests, RadialMR regression^56^ was run to detect SNP outliers. Outlier SNPs were removed. (A different number of telomere length SNPs were used for the various blood traits due to outliers being removed and whether a SNP or its “proxy” was available in the outcome dataset.) All instrumental variables included in this analysis have Cochrane’s *Q*-statistic *P*-values indicating no evidence for heterogeneity between SNPs^57^ (heterogeneity statistics are provided in Supplementary Tables 10–18).

The selected SNPs correspond to independent genomic regions and account for 1% to 2% of the variance in leukocyte telomere length (R^2^), which corresponds to *F*-statistics between 14 to 17. *F*-statistics are used to gauge whether the IVW results suffer from reduced statistical power to reject the null hypothesis. This could happen if telomere-length instrument explained a limited proportion of the variance in leukocyte telomere length. *F*-statistics > 10 are conventionally considered to be sutiable^58,59^. The *F*-statistics and R^2^ values used to calculate them are presented in Table 1.

**Table 1.**
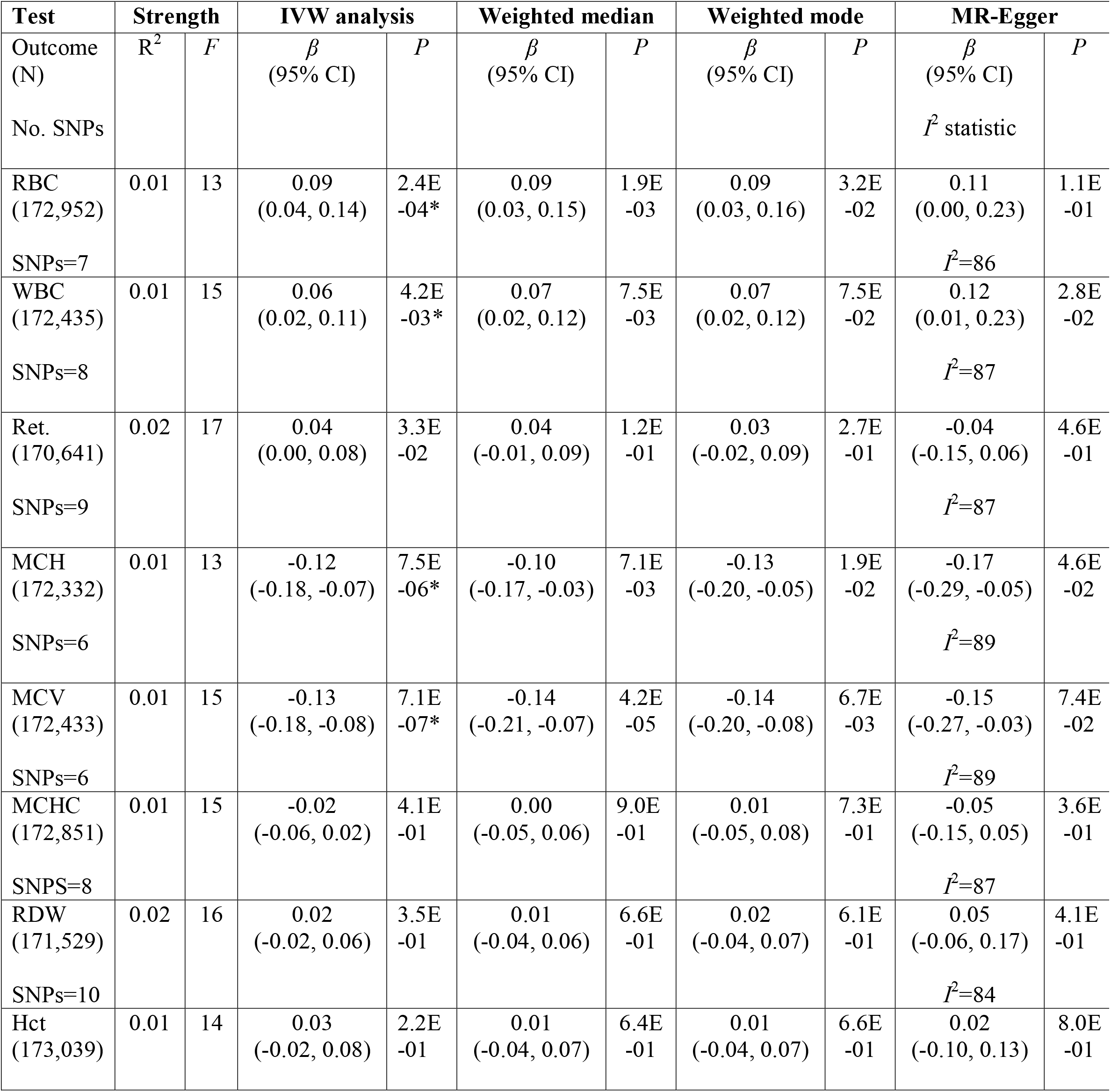
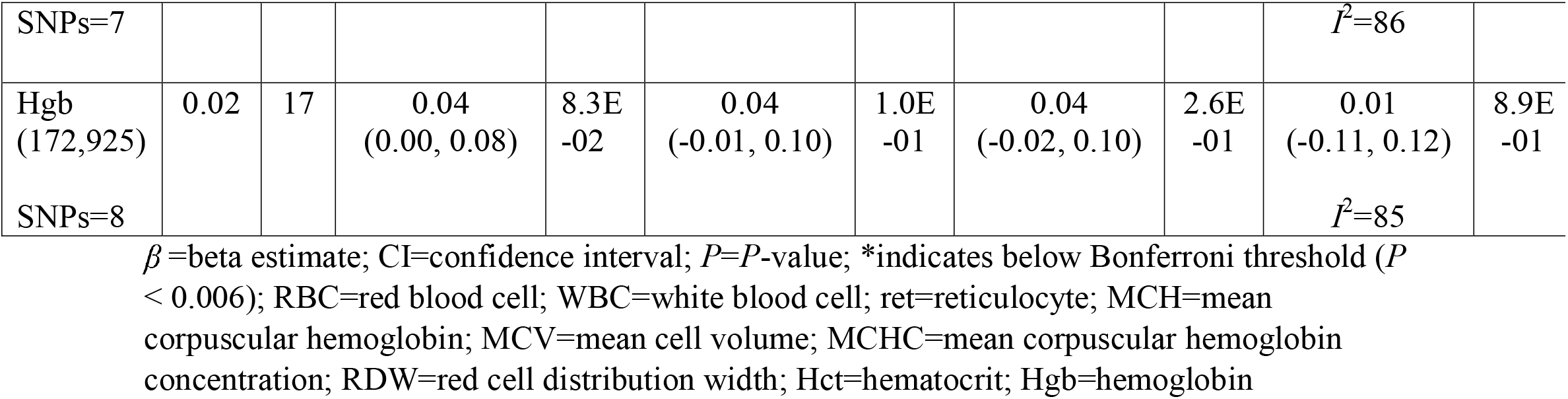
Estimates of the effect of genetically influenced longer telomere length on nine blood-cell traits.

In addition, *I*^2^ statistics, which are useful for assessing potential attenuation bias in one of the sensitivity estimators (the MR-Egger regression) are provided (also in Table 1). *I*^2^ statistics <90% can indicate dilution in the MR-Egger estimate, which can mean that the results from the MR-Egger intercept test (see below) are potentially inaccurate. Simulation extrapolation (SIMEX), a correction procedure that adjusts the MR-Egger estimate for potential regression dilution to the null, is recommended for *I*^2^ statistics <90%^60^. The SIMEX results are reported in Supplementary Tables 10–19.

### Sensitivity analyses

The IVW estimator can be biased if any of the instrumental SNPs violate the MR assumption about the genetic instrument not having a direct effect on the outcome independent of the exposure^61^. To assess possible violations to MR assumption (iii), three sensitivity estimators—MR-Egger regression, weighted median, and weighted mode methods— were run and their results compared with those of the IVW. The sensitivity estimators make different assumptions about the underlying nature of pleiotropy. Thus, if the directions and magnitudes of their effects comport with those of the IVW, this is a qualitative screen against pleiotropy (likewise, heterogeneity in their effects suggests pleiotropy)^62^. Comparing the IVW and sensitivity estimators is a form of triangulation and knowledge synthesis—i.e., the judgement process for whether the results are consistent with causality is like what investigators do when they perform a systematic review of studies with different methods, strenghts, and limitations.

In-depth explanations of the MR estimators and their assumptions have been covered elsewhere^61,63,64^. The results for the IVW and sensitivity estimators are reported in Table 1, except for the results of the MR-Egger intercept tests. MR-Egger regression provides both an effect estimate and a test for directional pleiotropy (the MR-Egger intercept test). The MR-Egger intercept test for pleiotropy is interpretted differently than the estimators providing tests for associations between telomere length and the blood-cell traits. When the MR-Egger intercept does not differ from zero (*P* > 0.05), this is evidence against pleiotropy. The MR-Egger intercept results are reported in Supplementary Tables 10–18.

### Number of tests

In total, nine MR tests were run (detailed characteristics for the individual SNPs used in each model are provided in Supplementary Tables 1–9). Though it is over conservative due to the correlation between blood traits, to account for multiple testing across analyses, a Bonferroni correction was used to establish a *P*-value threshold for strong evidence (*P* < 0.006) (false-positive rate = 0.05/9 outcomes).

### Statistical software

SIMEX corrections were perfomed in Stata SE/16.0^65^. All other described analyses were performed in R version 3.5.2 with the “TwoSampleMR” package^42^.

### Data availability

All data sources used for SNP-exposure and SNP-outcome associations are publicly available. The summary data for the telomere length instruments are available in Haycock et al. (2017)^20^. The nine hematological outcome GWA studies used for these analyses are accessible within MR-Base: http://www.mrbase.org/^42^.

## Data Availability

Data availability. All data sources used for SNP-exposure and SNP-outcome associations are publicly available. The summary data for the telomere length instruments are available in Haycock et al. (2017)20. The nine hematological outcome GWA studies used for these analyses are accessible within MR-Base: http://www.mrbase.org/.

## Abbreviations

MR: Mendelian randomization
RBC: red blood cell
WBC: white blood cell
Ret: reticulocyte
MCH: mean corpuscular hemoglobin
MCV: mean cell (corpuscular) volume
MCHC: mean corpuscular hemoglobin concentration
RDW: red blood cell distribution width
Hct: hematocrit
Hgb: hemoglobin
GWA: genome-wide association
HSC: hematopoietic stem cell
BMF: bone marrow failure
IBMFS: inherited bone marrow failure syndromes

